# Pre-dialysis Systolic Blood Pressure Time in Target Range and Risk of Stroke and Cardiovascular Events in Hemodialysis

**DOI:** 10.64898/2026.11.25.26369014

**Authors:** Fan-Yu Chen, Mu-Yang Hsieh, Zih-Kai Kao, Shao-Yuan Chuang, Ru-Yin Shu, Chen-Huan Chen, Mei-Jy Jeng, Chih-Cheng Wu, Hao-Min Cheng

## Abstract

**Background:** Patients receiving maintenance hemodialysis are at increased risk of cerebrovascular events, but conventional blood pressure metrics may not adequately capture longitudinal hemodynamic stability. We examined whether pre-dialysis systolic blood pressure time in target range (SBP-TTR) was associated with stroke and cardiovascular outcomes.

**Methods:** In this prospective multicenter cohort, 1,136 adults receiving thrice-weekly hemodialysis underwent a 12-week baseline assessment with up to 36 pre-dialysis blood pressure measurements. Restricted cubic splines evaluated associations between mean systolic blood pressure and clinical outcomes. SBP-TTR within 130–150 mmHg was calculated using Rosendaal linear interpolation. The primary outcome was major adverse cardiovascular events (MACE), comprising nonfatal stroke, cardiovascular death, and nonfatal myocardial infarction. Multivariable Cox and Fine–Gray competing-risk models adjusted for clinical characteristics, dialysis-related factors, medications, mean systolic blood pressure, and blood pressure variability.

**Results:** Mean pre-dialysis systolic blood pressure showed U-shaped associations with MACE and nonfatal stroke, with risk nadirs of 141.3 and 138.2 mmHg, respectively. Each 10-percentage-point increase in SBP-TTR was associated with lower risks of MACE (hazard ratio, 0.93 [95% CI, 0.88–0.98]; P=0.005) and nonfatal stroke (hazard ratio, 0.89 [95% CI, 0.83–0.96]; P=0.003), but not cardiovascular death or nonfatal myocardial infarction. Associations remained consistent in competing-risk analyses. SBP-TTR improved risk discrimination and reclassification beyond conventional blood pressure measures.

**Conclusions:** Greater pre-dialysis SBP-TTR within 130–150 mmHg was associated with lower risks of nonfatal stroke and MACE, independent of mean blood pressure and blood pressure variability. SBP-TTR may provide complementary information for cerebrovascular and cardiovascular risk assessment in hemodialysis.

## Introduction

Cardiovascular disease (CVD) remains the leading cause of morbidity and mortality in patients with end-stage renal disease (ESRD) receiving maintenance hemodialysis (MHD).^1^ Although contemporary ESC and ACC/AHA guidelines advocate intensive blood pressure (BP) control,^2,3^ BP management in hemodialysis (HD) remains controversial. The 2021 KDIGO guideline recommends a systolic blood pressure (SBP) target of <120 mmHg in non-dialysis (ND) chronic kidney disease,^4^ yet this threshold has not been validated in HD. In HD, pre-dialysis SBP is strongly influenced by volume status, interdialytic fluid accumulation, ultrafiltration (UF), arterial stiffness, and autonomic dysfunction, resulting in substantial fluctuations both across and within sessions. Consequently, observational studies report paradoxical U-shaped associations between SBP and adverse outcomes in dialysis cohorts,^5,6^ suggesting that both low and high pre-dialysis SBP reflect distinct pathophysiological states. Therefore, defining a ideal pre-dialysis SBP range in HD remains a major challenge.

Current BP assessment in HD relies primarily on mean SBP and visit-to-visit BP variability.^7,8^ Mean SBP reflects overall exposure but not temporal control, whereas variability indices capture dispersion without accounting for time spent within target ranges. Time in target range (TTR) addresses this limitation by quantifying the proportion of time BP remains within a predefined range, providing a longitudinal measure that integrates BP level and stability over time.^9^ In ND populations, higher SBP-TTR is associated with lower risks of mortality and major adverse cardiovascular events (MACE), independent of mean SBP and variability,^10^ with similar findings in essential hypertension, heart failure (HF), and incident atrial fibrillation (AF).^11–13^ However, the meaning of SBP-TTR in HD cannot be assumed to be equivalent to that in ND populations. In HD, repeated pre-dialysis SBP measurements are embedded within cyclical changes in extracellular volume, dry-weight management, UF exposure and tolerance, and antihypertensive treatment practices. Thus, SBP-TTR may reflect not only pharmacological BP control but also dialysis-specific longitudinal hemodynamic phenotype. Low SBP-TTR may reflect recurrent above-range SBP related to volume overload, below-range SBP associated with excessive BP lowering, or limited cardiovascular reserve, or greater fluctuations arising from UF intolerance, frailty, or poor intake. Whether SBP-TTR retains prognostic significance beyond mean SBP and BP variability in this physiologically distinct setting remains uncertain.

We therefore conducted a prospective multicenter cohort study to identify a clinically relevant pre-dialysis SBP range, quantify SBP-TTR using time-weighted linear interpolation, evaluate its association with cardiovascular (CV) outcomes independent of mean SBP and BP variability, and assess its incremental prognostic value beyond these conventional BP metrics.

## Methods

### Data Availability Statement

The individual participant data underlying this article cannot be made publicly available because participants did not provide consent for unrestricted data sharing and because of institutional and data protection requirements. Deidentified participant data and the data dictionary may be made available to qualified researchers upon reasonable request to the corresponding author, subject to approval by the relevant institutional review board and execution of an appropriate data-use agreement. The analytic code used to derive SBP-TTR and reproduce the principal analysis will be deposited in a public repository, and a persistent link will be provided before publication.

### Study design, data source, and population

The Hsinchu Vascular Access (VA) Study (ClinicalTrials.gov identifier NCT04692636) is a multicenter, prospective observational cohort of adults receiving MHD designed to identify determinants of VA and CV events. Between 1 January 2018 and 30 June 2019, consecutive patients were screened across 12 dialysis units in Hsinchu, Taiwan (4 hospital-based and 8 standalone facilities). Clinical, dialysis-related, and laboratory data were prospectively extracted from electronic records using prespecified, harmonized definitions. A 12-week baseline period characterized longitudinal BP exposure and reduced the influence of short-term hemodynamic variability.

Eligible participants were 18–90 years old, received thrice-weekly HD for at least 6 months, and had not been hospitalized within 3 months. Exclusion criteria were death, transfer, dialysis-modality change, or incomplete baseline BP data (<30 of 36 scheduled pre-dialysis measurements). Follow-up began after baseline and continued through 31 December 2022. Participants were censored at death, kidney transplantation, transfer to other study facility, or administrative end of follow-up. All censoring events were recorded, and no participants were lost to follow-up. Outcomes, covariates, candidate SBP ranges, and analytical procedures were prespecified. The primary SBP range was subsequently selected from the prespecified candidate ranges based on the observed associations with cardiovascular outcomes.

### BP measurement and data collection

Pre-dialysis BP was measured during routine outpatient care using a standardized protocol and a validated automated oscillometric device (Omron HBP-1300 Professional BP Monitor; Omron Healthcare, Kyoto, Japan) on the arm contralateral to the VA. Devices underwent routine calibration and performance checks. BP was recorded before cannulation after at least 5 minutes of seated rest, with the back supported, feet flat, and arm at heart level. Two readings obtained 1–2 minutes apart were averaged and automatically transmitted to a centralized dialysis information system. The 12-week baseline included 36 planned sessions per participant; all available pre-dialysis measurements were analyzed.

### Definition of SBP-TTR

SBP time in target range (SBP-TTR) was defined a priori as a longitudinal, time-weighted measure of pre-dialysis SBP stability during baseline. The primary definition used the Rosendaal linear interpolation method,^14^ assuming linear change between consecutive measurements, to estimate the percentage of observation time within the selected target range. The range was empirically derived from CV outcome associations as described below (**Definition of target BP ranges**). Interpolation accounted for irregular measurement intervals and reflected cumulative BP burden across dialysis sessions. SBP-TTR was expressed as a percentage of total observation time and reflected both cumulative BP burden and stability across repeated dialysis sessions.

Two alternative calculation methods were used in sensitivity analysis. Proportion-based SBP-TTR was the percentage of discrete pre-dialysis SBP readings within range without accounting for time between measurements. Dual-target SBP × diastolic BP (DBP)-TTR estimated the percentage of time both measures were simultaneously within their respective ranges, testing whether combined hemodynamic control added information beyond SBP alone. All TTR measures were analyzed continuously per 10-percentage-point increase and categorically according to their population-specific quartiles. The calculation methods are illustrated in **Supplementary Figure S1**.

### Definition of target BP ranges

Because no guideline-endorsed pre-dialysis SBP range exists for patients receiving MHD, we evaluated 12 clinically interpretable candidate ranges of different widths. These included four 10-mmHg ranges (120–130, 130–140, 140–150, and 150–160 mmHg), four 20-mmHg ranges (110–130, 120–140, 130–150, and 140–160 mmHg), and four 30-mmHg ranges (100–130, 110–140, 120–150, and 130–160 mmHg). Mean pre-dialysis SBP was first examined using restricted cubic splines in Cox proportional hazards models for MACE and its individual components, with knots placed at the 10th, 50th, and 95th percentiles. Associations between SBP-TTR and cardiovascular outcomes were then evaluated across all candidate ranges. The range used for subsequent detailed analysis was selected according to its alignment with the spline-derived lowest-risk region, consistency of associations across CV outcomes, and clinical interpretability. The rationale for selecting the final primary range is reported in the **Results**.

### Clinical outcomes

Participants were reviewed at standardized 3-month intervals through dialysis records to ascertain vital status and interim events. Trained facility coordinators recorded event dates, which principal investigators adjudicated through independent medical-record review and, when needed, consultation with treating physicians. Outcomes were prespecified. The primary outcome was MACE, defined as the first occurrence of nonfatal stroke, CV death, or nonfatal myocardial infarction (MI). Secondary outcomes were the individual components. Exploratory outcomes included all-cause mortality, incident CVD, and cerebrovascular outcomes. Incident CVD comprised MACE, percutaneous coronary intervention, and chronic coronary syndrome. Stroke was classified as ischemic or hemorrhagic; ischemic subtypes were categorized as large-artery atherosclerosis, small-vessel disease, or cardioembolism using the Trial of Org 10172 in Acute Stroke Treatment classification.

### Covariates

Prespecified covariates were obtained during baseline. Missing information was supplemented through clinical visits or telephone monitoring, and medical history was obtained by structured interview. Baseline characteristics included age, sex, body mass index, smoking, hypertension, diabetes mellitus, dyslipidemia, CVD, coronary artery disease, HF, peripheral artery disease, cerebrovascular disease, and AF. Dialysis covariates included vintage, adequacy assessed by single-pool Kt/V, interdialytic weight gain (IDWG), and UF volume expressed as a percentage of dry weight. IDWG was the difference between pre-dialysis weight and the preceding postdialysis weight; both weight gain and UF volume were averaged across baseline. Laboratory covariates included albumin, hemoglobin, total cholesterol, calcium, phosphorus, and calcium–phosphorus product. Medications included antiplatelet agents, vitamin K antagonists, lipid-lowering agents, β-blockers, renin– angiotensin system inhibitors, and calcium channel blockers. BP covariates included baseline and mean systolic and diastolic BP, mean arterial pressure, and visit-to-visit variability measured by standard deviation (SD), coefficient of variation, and average real variability (ARV). Mean arterial pressure was calculated as DBP + 1/3(SBP − DBP).

### Statistical analysis

Continuous variables are presented as mean (SD) and categorical variables as counts (percentages). Groups were compared using analysis of variance or Kruskal–Wallis tests for continuous variables and χ² or Fisher exact tests for categorical variables, as appropriate. Mean pre-dialysis SBP was first examined using restricted cubic splines within Cox proportional hazards regression to identify the lowest-risk region for MACE. Based on the nadir and curve shape, a clinically applicable 20-mmHg range was defined around this region, balancing interpretability and measurement variability. SBP-TTR was then analyzed continuously per 10-percentage-point increase and categorically by quartiles. Overall and nonlinearity tests were reported for spline models.

Cox proportional hazards models estimated hazard ratios (HR) and 95% confidence intervals (CI) for MACE, all-cause mortality, and other prespecified outcomes; proportional hazards assumptions were evaluated using Schoenfeld residuals. Fine–Gray models estimated subdistribution hazard ratios (SHR), with all-cause death as the competing event for nonfatal outcomes and noncardiovascular death for CV death. Follow-up began after completion of baseline. Kaplan–Meier curves illustrated event-free survival and were compared using log-rank tests. Cumulative incidence functions were used when competing risks were present and compared using Gray tests. Primary adjustment included sex, age, smoking, diabetes, CV and cerebrovascular disease, dialysis vintage and adequacy, IDWG, UF volume, albumin, and baseline medication use. Sequential models additionally incorporated mean SBP, SBP SD, or both.

Multivariable restricted cubic splines characterized continuous associations between SBP-TTR and outcomes across the full exposure distribution. Sensitivity analysis examined alternative target ranges and TTR definitions. Prespecified subgroup analysis assessed effect modification across clinically relevant strata, including sex, age, body mass index, smoking, baseline systolic and diastolic BP, hypertension, diabetes, and dyslipidemia. Sensitivity analysis also included categorical TTR analysis and Fine–Gray models using the alternative TTR definitions.

To assess independent hemodynamic contributions, a clinical reference model was followed by sequential addition of SBP-TTR, mean SBP, and SBP variability. Correlations among BP metrics were examined (**Supplementary Figure S5**). To reduce collinearity, residual components of mean SBP and BP variability were derived by linear regression and included in combined models (**Supplementary Figure S6**), assuming linear relationships among BP metrics. Detailed model specifications and refinement procedures are provided in **Supplementary Table S18**. Incremental prognostic value was evaluated using changes in −2 log likelihood, Harrell’s C-index, net reclassification improvement (NRI), and integrated discrimination improvement (IDI).

Benjamini–Hochberg false discovery rate (FDR) adjustment addressed multiple comparisons across outcomes. All tests were two-sided, with P < 0.05 considered statistically significant. Analysis used R version 4.5.0 (R Foundation for Statistical Computing, Vienna, Austria); survival supported Cox regression, survminer and adjustedCurves generated survival curves, ggsurvfit and cmprsk supported cumulative-incidence and competing-risk analysis, and survIDINRI calculated reclassification metrics.

### Ethical approval

The Institutional Review Board of Hsinchu Veterans General Hospital approved the study (HCH-109-089-E), which was conducted in accordance with the Declaration of Helsinki. All participants provided written informed consent before enrollment.

## Results

### Identification of a clinically relevant pre-dialysis SBP range

Of 1,215 patients screened, 30 were excluded because of dialysis duration <6 months or recent hospitalization, and 49 were excluded during the 12-week baseline period because of death, transfer, modality change, or incomplete BP data, yielding 1,136 participants (**Figure 1**). Mean pre-dialysis SBP showed U-shaped associations with MACE and nonfatal stroke (**Figure 2**). Estimated risk nadirs were 141.3 mmHg for MACE (*P* for overall association = 0.017; *P* for nonlinearity = 0.004) and 138.2 mmHg for nonfatal stroke (*P* for overall association = 0.003; *P* for nonlinearity = 0.003). Associations with CV death and nonfatal MI were not statistically significant. The 130–150 mmHg range, centered on the MACE risk nadir, was selected for the primary SBP-TTR analysis. Across prespecified 10-, 20-, and 30-mmHg windows, this range showed the most consistent associations with lower risks of MACE and nonfatal stroke, whereas associations with CV death and nonfatal MI were less consistent (**Figure 3**). Within the selected range, each 10-percentage-point increase in interpolation-based SBP-TTR was associated with lower MACE risk (HR, 0.93; 95% CI, 0.88–0.98; *P* = 0.005). A stricter dual-target definition requiring SBP of 130–150 mmHg and DBP of 70–90 mmHg produced a similar estimate (HR, 0.91; 95% CI, 0.84–0.99; *P* = 0.026).

**Figure 1.**
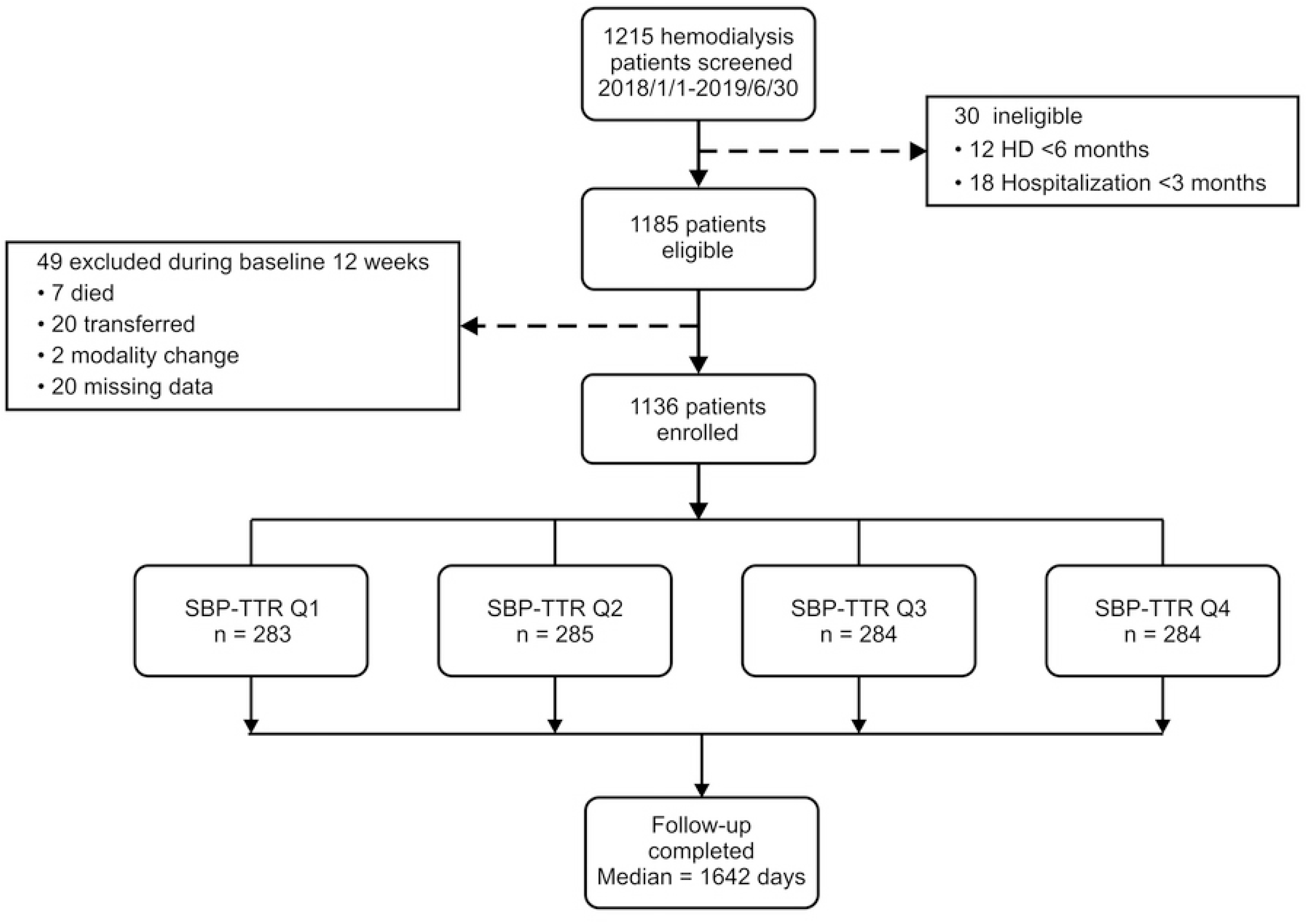
Selection of the Study Cohort. The flow diagram shows the selection of patients receiving MHD for inclusion in the analytic cohort. SBP-TTR was calculated during the 12-week baseline period using linear interpolation of routinely collected pre-dialysis SBP measurements. MHD, maintenance hemodialysis; SBP-TTR, systolic blood pressure time in target range. Alt text: Flow diagram showing the selection of 1,136 patients from 1,215 screened after application of eligibility criteria and baseline exclusions.

**Figure 2.**
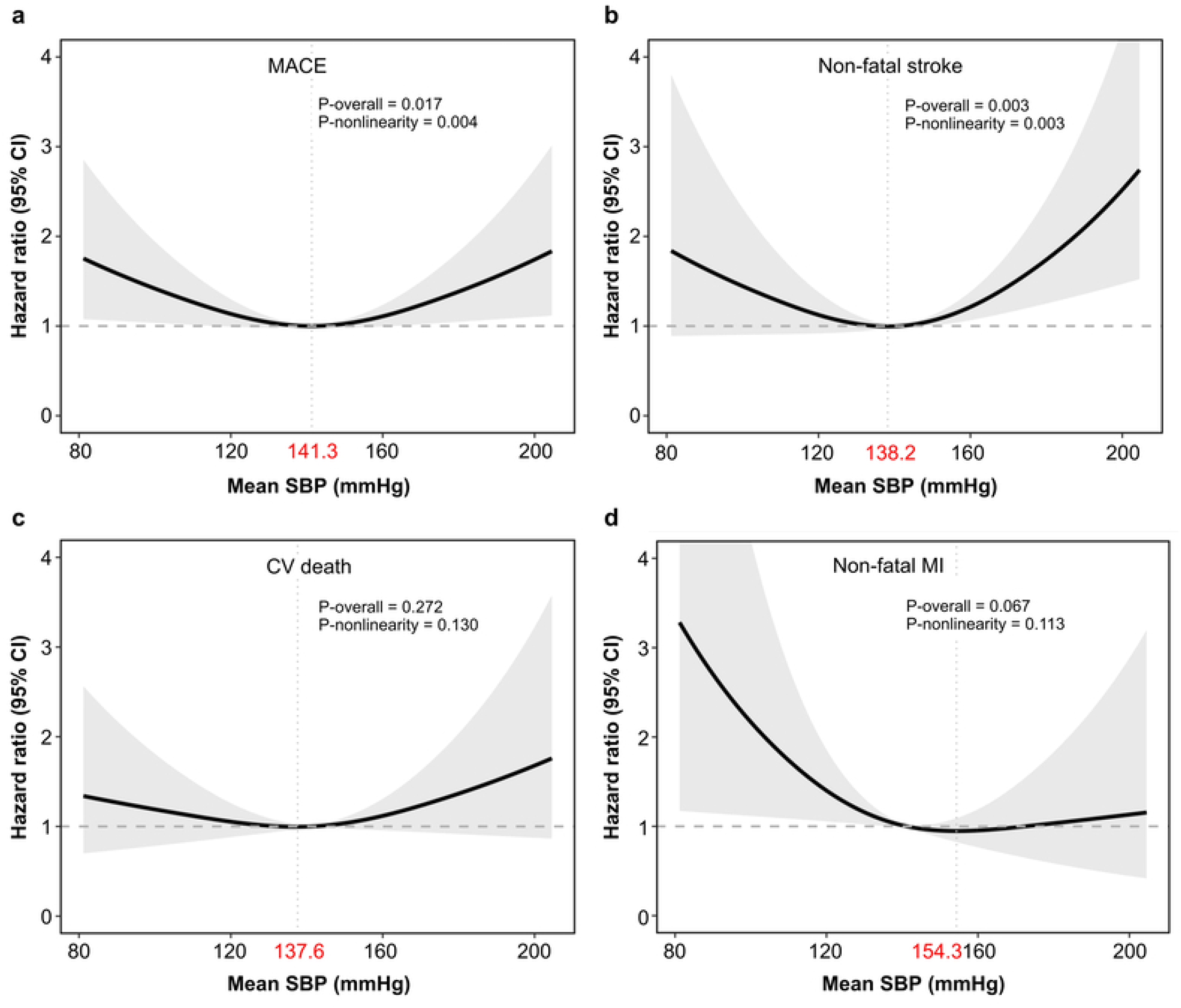
Associations Between Mean Pre-dialysis SBP and Cardiovascular Outcomes. Restricted cubic spline models evaluated the associations of mean pre-dialysis SBP during the 12-week baseline period with MACE (A), nonfatal stroke (B), CV death (C), and nonfatal MI (D). Solid lines represent adjusted HRs, shaded areas represent 95% CIs, horizontal dashed lines indicate an HR of 1.0, and vertical dotted lines indicate the estimated risk nadirs. Models were adjusted for sex, age, smoking status, diabetes mellitus, CVD, cerebrovascular disease, dialysis vintage, dialysis adequacy, IDWG, UF volume, serum albumin, medication use, and SBP SD. P-overall indicates the overall association, and P-nonlinearity indicates departure from linearity. CI, confidence interval; CV, cardiovascular; CVD, cardiovascular disease; HR, hazard ratio; IDWG, interdialytic weight gain; MACE, major adverse cardiovascular events; MI, myocardial infarction; SBP, systolic blood pressure; SD, standard deviation; UF, ultrafiltration. Alt text: Four restricted cubic spline plots showing U-shaped associations of mean pre-dialysis SBP with MACE and nonfatal stroke, with estimated risk nadirs near 141 and 138 mmHg, respectively. Associations with CV death and nonfatal MI were not statistically significant.

**Figure 3.**
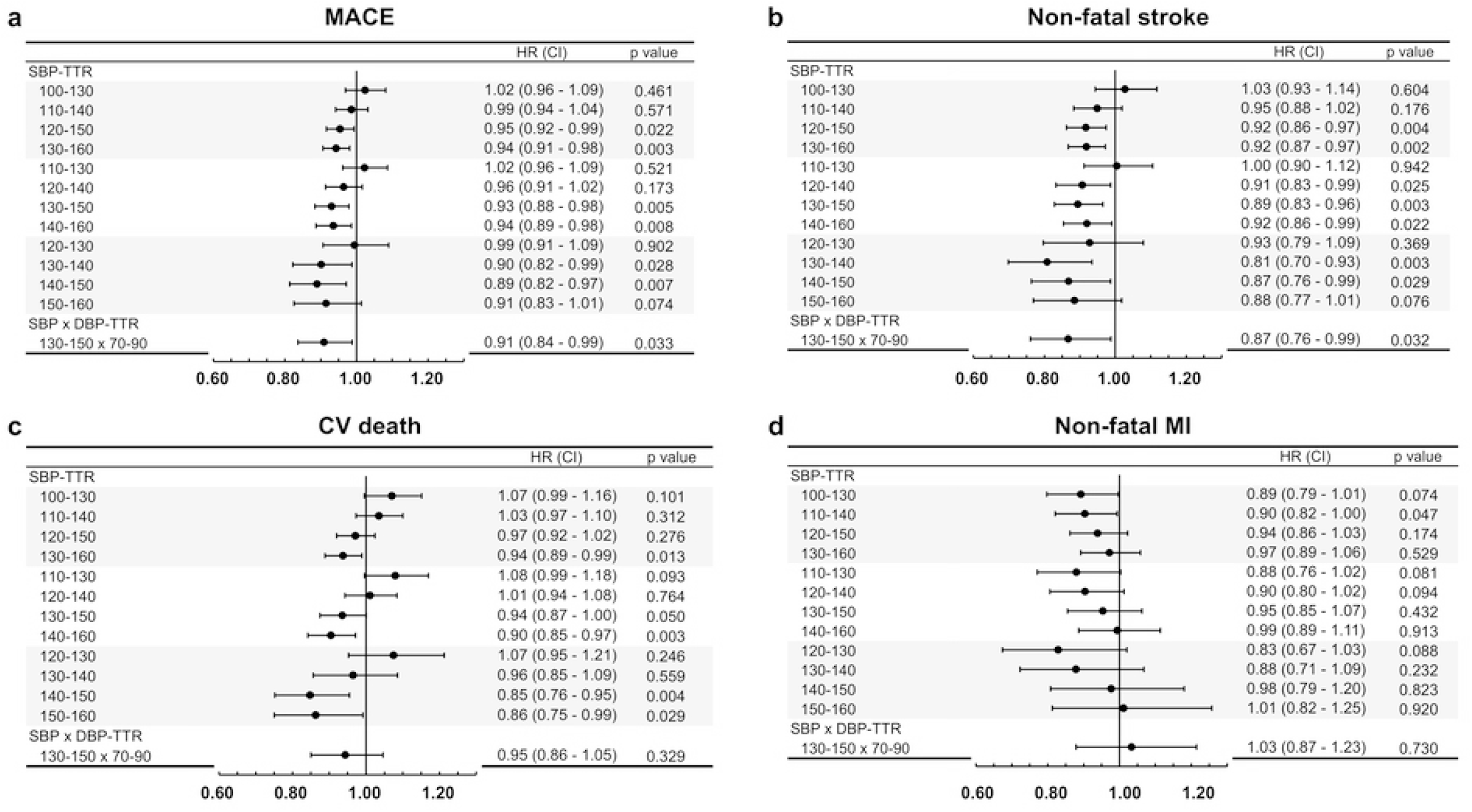
Associations Between SBP-TTR and Cardiovascular Outcomes Across Candidate Target Ranges. HRs are shown per 10-percentage-point increase in SBP-TTR across prespecified candidate SBP ranges for MACE (A), nonfatal stroke (B), CV death (C), and nonfatal MI (D). SBP-TTR was calculated during the 12-week baseline period using linear interpolation. Candidate ranges comprised overlapping 10-, 20-, and 30-mmHg windows; dual-target SBP×DBP TTR required simultaneous pre-dialysis SBP of 130–150 mmHg and DBP of 70–90 mmHg. Points represent HRs, horizontal lines represent 95% CIs, and the vertical line indicates an HR of 1.0. Models were adjusted for sex, age, smoking status, diabetes mellitus, CVD, cerebrovascular disease, dialysis vintage, dialysis adequacy, IDWG, UF volume, serum albumin, medication use, mean SBP, and SBP SD. CI, confidence interval; CV, cardiovascular; CVD, cardiovascular disease; DBP, diastolic blood pressure; HR, hazard ratio; IDWG, interdialytic weight gain; MACE, major adverse cardiovascular events; MI, myocardial infarction; SBP-TTR, systolic blood pressure time in target range; SD, standard deviation; UF, ultrafiltration. Alt text: Forest plots comparing associations of SBP-TTR with CV outcomes across candidate SBP ranges. The 130–150 mmHg range showed the most consistent inverse associations with MACE and nonfatal stroke, whereas associations with CV death and nonfatal MI were less consistent.

### Study population and baseline characteristics

Participants were categorized into quartiles of interpolation-based SBP-TTR: 0% to <7.4% (n = 283), 7.4% to <25.9% (n = 285), 25.9% to <45.9% (n = 284), and 45.9% to 100% (n = 284) (**Figure 1**). Mean age was 66.7 years, and 52.1% were male. Major comorbidities, dialysis adequacy, and laboratory parameters were generally similar across quartiles (**Table 1**). Compared with the lowest quartile, participants in the highest quartile were older, had shorter dialysis vintage, and more frequently received lipid-lowering agents; use of other medications was similar. Despite marked differences in SBP-TTR, mean pre-dialysis SBP was nearly identical across quartiles (overall mean, 141.2 mmHg; *P* = 0.958). In contrast, higher SBP-TTR was associated with lower visit-to-visit SBP SD, coefficient of variation, and ARV (all *P* < 0.001; **Supplementary Table S1**).

**Table 1.** Baseline Characteristics According to Quartiles of Pre-dialysis SBP Time in Target Range.

|  | Total participants | Q1 | Q2 | Q3 | Q4 |  |
|---|---|---|---|---|---|---|
|  | (n = 1,136) | 0% to <7.4%<br>(n = 283) | 7.4% to <25.9%<br>(n = 285) | 25.9% to <45.9%<br>(n = 284) | 45.9% to 100%<br>(n = 284) | P value |
| Male, n (%) | 592 (52.1) | 135 (47.7) | 167 (58.6) | 148 (52.1) | 142 (50.0) | 0.057 |
| Age, years | 66.7 ± 13.9 | 64.6 ± 14.0 | 66.5 ± 14.4 | 67.0 ± 13.8 | 68.5 ± 12.9 | 0.009 |
| Body mass index, kg/m² | 22.9 ± 4.2 | 22.5 ± 4.1 | 23.0 ± 4.2 | 23.3 ± 4.4 | 22.6 ± 4.0 | 0.101 |
| Current smoking, n (%) | 164 (14.4) | 42 (14.8) | 32 (11.2) | 48 (16.9) | 42 (14.8) | 0.279 |
| Diabetes mellitus, n (%) | 608 (53.5) | 137 (48.4) | 161 (56.5) | 152 (53.5) | 158 (55.6) | 0.213 |
| Dyslipidemia, n (%) | 234 (20.6) | 59 (20.8) | 52 (18.2) | 63 (22.2) | 60 (21.1) | 0.692 |
| Cardiovascular disease, n (%) | 440 (38.7) | 96 (33.9) | 114 (40.0) | 114 (40.1) | 116 (40.8) | 0.293 |
| Coronary artery disease, n (%) | 284 (25.0) | 67 (23.7) | 72 (25.3) | 72 (25.4) | 73 (25.7) | 0.946 |
| Heart failure, n (%) | 134 (11.8) | 27 (9.5) | 33 (11.6) | 36 (12.7) | 38 (13.4) | 0.514 |
| Peripheral artery disease, n (%) | 99 (8.7) | 16 (5.7) | 29 (10.2) | 26 (9.2) | 28 (9.9) | 0.201 |
| Cerebrovascular disease, n (%) | 100 (8.8) | 23 (8.1) | 30 (10.5) | 24 (8.5) | 23 (8.1) | 0.697 |
| Atrial fibrillation, n (%) | 173 (15.2) | 39 (13.8) | 51 (17.9) | 40 (14.1) | 43 (15.1) | 0.509 |
| Dialysis vintage, years | 4.6 ± 5.3 | 5.4 ± 5.8 | 4.4 ± 5.1 | 4.2 ± 4.7 | 4.4 ± 5.3 | 0.032 |
| Dialysis adequacy, spKt/V | 1.7 ± 0.3 | 1.7 ± 0.3 | 1.7 ± 0.2 | 1.7 ± 0.3 | 1.7 ± 0.4 | 0.723 |
| Interdialytic weight gain, kg | 2.2 ± 1.0 | 2.3 ± 1.0 | 2.2 ± 1.1 | 2.2 ± 1.0 | 2.1 ± 0.9 | 0.238 |
| Ultrafiltration volume, % of dry weight | 3.6 ± 1.4 | 3.8 ± 1.4 | 3.6 ± 1.6 | 3.5 ± 1.3 | 3.5 ± 1.3 | 0.061 |
| Serum albumin, g/dL | 3.8 ± 0.3 | 3.8 ± 0.3 | 3.8 ± 0.3 | 3.8 ± 0.3 | 3.8 ± 0.3 | 0.903 |
| Hemoglobin, g/dL | 10.7 ± 1.3 | 10.7 ± 1.4 | 10.7 ± 1.3 | 10.6 ± 1.3 | 10.6 ± 1.4 | 0.527 |
| Total cholesterol, mg/dL | 159.9 ± 30.5 | 162.6 ± 29.6 | 157.6 ± 28.1 | 160.2 ± 34.2 | 159.1 ± 29.8 | 0.193 |
| Serum calcium, mg/dL | 9.3 ± 0.9 | 9.2 ± 0.8 | 9.4 ± 0.8 | 9.3 ± 0.9 | 9.3 ± 0.9 | 0.078 |
| Serum phosphorus, mg/dL | 5.0 ± 1.4 | 5.0 ± 1.5 | 4.9 ± 1.3 | 5.0 ± 1.4 | 4.9 ± 1.3 | 0.470 |
| Calcium–phosphorus product, mg²/dL² | 52.8 ± 15.8 | 53.7 ± 17.1 | 52.5 ± 15.0 | 53.3 ± 15.2 | 51.7 ± 16.0 | 0.557 |
| Antiplatelet therapy, n (%) | 279 (24.6) | 65 (23.0) | 62 (21.8) | 71 (25.0) | 81 (28.5) | 0.258 |
| Vitamin K antagonist, n (%) | 11 (1.0) | 6 (2.1) | 2 (0.7) | 0 (0.0) | 3 (1.1) | 0.055 |
| Lipid-lowering agents, n (%) | 147 (12.9) | 17 (6.0) | 39 (13.7) | 48 (16.9) | 43 (15.1) | <0.001 |
| β-Blockers, n (%) | 195 (17.2) | 47 (16.6) | 52 (18.2) | 46 (16.2) | 50 (17.6) | 0.914 |
| Renin–angiotensin system inhibitors, n (%) | 189 (16.6) | 41 (14.5) | 46 (16.1) | 45 (15.8) | 57 (20.1) | 0.316 |
| Calcium channel blockers, n (%) | 279 (24.6) | 57 (20.1) | 73 (25.6) | 67 (23.6) | 82 (28.9) | 0.105 |
| Number of antihypertensive agents |  |  |  |  |  | 0.147 |
| 0 | 752 (66.2) | 205 (72.4) | 186 (65.3) | 185 (65.1) | 176 (62.0) |  |
| 1 | 173 (15.2) | 28 (9.9) | 46 (16.1) | 52 (18.3) | 47 (16.5) |  |
| 2 | 143 (12.6) | 33 (11.7) | 34 (11.9) | 35 (12.3) | 41 (14.4) |  |
| 3 | 68 (6.0) | 17 (6.0) | 19 (6.7) | 12 (4.2) | 20 (7.0) |  |

Data are mean ± standard deviation or number with percentage. SBP-TTR was defined as the percentage of interpolated time during the 12-week baseline period in which pre-dialysis SBP was within 130–150 mmHg. Quartiles were defined according to the distribution of interpolation-based SBP-TTR in the study population. SBP-TTR, systolic blood pressure time in target range; spKt/V, single-pool Kt/V.

### Time-to-event analysis and associations with clinical outcomes

Adjusted cumulative incidence curves showed lower cumulative incidences of MACE and nonfatal stroke among participants with higher SBP-TTR, whereas separation was less apparent for CV death and nonfatal MI (**Figure 4**). Kaplan–Meier and adjusted survival curves showed similar patterns **(Supplementary Figures S2 and S3)**. In fully adjusted Cox models, each 10-percentage-point increase in SBP-TTR was associated with lower risks of MACE (HR, 0.93; 95% CI, 0.88–0.98; *P* = 0.005) and nonfatal stroke (HR, 0.89; 95% CI, 0.83–0.96; *P* = 0.003), but not CV death (HR, 0.94; 95% CI, 0.88–1.00; *P* = 0.050) or nonfatal MI (HR, 0.95; 95% CI, 0.85–1.07; *P* = 0.432) (**Table 2**). In exploratory analysis, higher SBP-TTR was associated with lower incident CVD (HR, 0.94; 95% CI, 0.90–0.99), ischemic stroke (HR, 0.90; 95% CI, 0.82–0.97), and large-artery atherosclerosis (HR, 0.86; 95% CI, 0.74–0.99), but not all-cause mortality **(Supplementary Table S2)**. After FDR adjustment, associations were most consistent for cerebrovascular outcomes **(Supplementary Tables S16 and S17)**. Fine–Gray competing-risk analysis yielded similar estimates for MACE (SHR, 0.93; 95% CI, 0.88–0.98; *P* = 0.005) and nonfatal stroke (SHR, 0.89; 95% CI, 0.83–0.96; *P* = 0.003), with no significant associations for CV death or nonfatal MI (**Table 3**).

**Figure 4.**
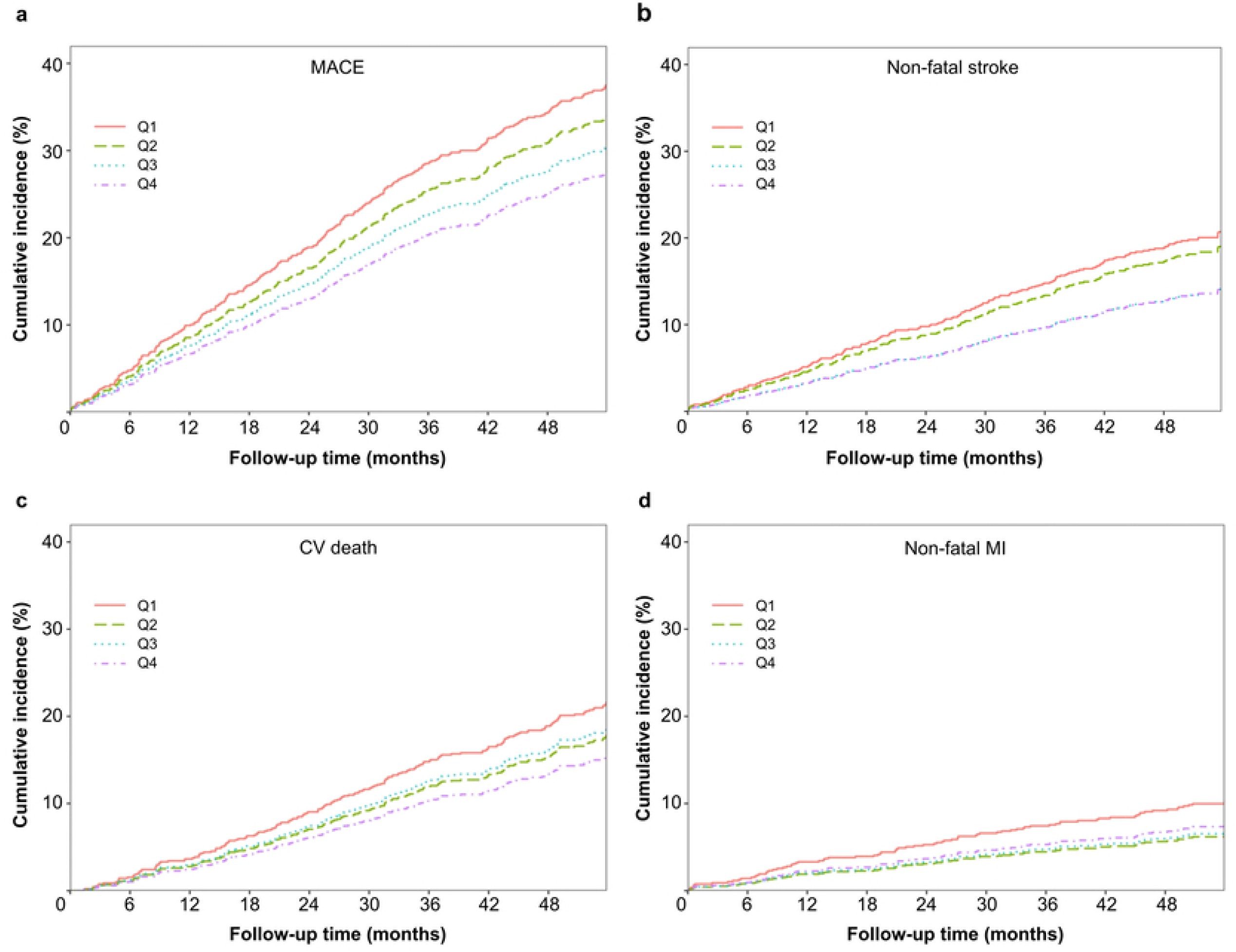
Adjusted Cumulative Incidence of Cardiovascular Outcomes According to SBP-TTR Quartiles. Adjusted cumulative incidence functions are shown for MACE (A), nonfatal stroke (B), CV death (C), and nonfatal MI (D) according to interpolation-based SBP-TTR quartiles. SBP-TTR was defined as the percentage of interpolated time during the 12-week baseline period in which pre-dialysis SBP was within 130–150 mmHg. All-cause death was treated as the competing event for nonfatal stroke and nonfatal MI, whereas non-CV death was treated as the competing event for CV death. Models were adjusted for sex, age, smoking status, diabetes mellitus, CVD, cerebrovascular disease, dialysis vintage, dialysis adequacy, IDWG, UF volume, serum albumin, medication use, mean SBP, and SBP SD. CV, cardiovascular; CVD, cardiovascular disease; IDWG, interdialytic weight gain; MACE, major adverse cardiovascular events; MI, myocardial infarction; SBP-TTR, systolic blood pressure time in target range; SD, standard deviation; UF, ultrafiltration. Alt text: Four adjusted cumulative incidence plots according to SBP-TTR quartiles. Separation across quartiles was more apparent for MACE and nonfatal stroke than for CV death or nonfatal MI.

**Table 2.** Associations Between SBP-TTR and Cardiovascular Outcomes in Cox Proportional Hazards Models.

|  | Events (n) | Incidence rate (per 100 person-years) | HR (95% CI) | P value |
|---|---|---|---|---|
| MACE | 350 | 9.29 |  |  |
| Crude |  |  | 0.95 (0.90 - 0.99) | 0.018 |
| Model 1 |  |  | 0.92 (0.88 - 0.97) | 0.001 |
| Model 2 |  |  | 0.92 (0.88 - 0.97) | 0.001 |
| Model 3 |  |  | 0.93 (0.88 - 0.98) | 0.006 |
| Model 4 |  |  | 0.93 (0.88 - 0.98) | 0.005 |
| Nonfatal stroke | 185 | 5.39 |  |  |
| Crude |  |  | 0.89 (0.83 - 0.95) | <0.001 |
| Model 1 |  |  | 0.86 (0.81 - 0.92) | <0.001 |
| Model 2 |  |  | 0.87 (0.82 - 0.93) | <0.001 |
| Model 3 |  |  | 0.89 (0.83 - 0.96) | 0.002 |
| Model 4 |  |  | 0.89 (0.83 - 0.96) | 0.003 |
| CV death | 193 | 4.80 |  |  |
| Crude |  |  | 0.96 (0.90 - 1.02) | 0.162 |
| Model 1 |  |  | 0.93 (0.88 - 1.00) | 0.043 |
| Model 2 |  |  | 0.93 (0.88 - 1.00) | 0.040 |
| Model 3 |  |  | 0.94 (0.88 - 1.00) | 0.059 |
| Model 4 |  |  | 0.94 (0.88 - 1.00) | 0.050 |
| Nonfatal MI | 83 | 2.33 |  |  |
| Crude |  |  | 0.96 (0.87 - 1.05) | 0.375 |
| Model 1 |  |  | 0.94 (0.84 - 1.05) | 0.239 |
| Model 2 |  |  | 0.93 (0.84 - 1.04) | 0.225 |
| Model 3 |  |  | 0.95 (0.85 - 1.07) | 0.440 |
| Model 4 |  |  | 0.95 (0.85 - 1.07) | 0.432 |

Incidence rates are per 100 person-years; HRs are per 10% increment in interpolation-based SBP-TTR. Model 1 included sex, age, smoking status, diabetes mellitus, cardiovascular disease, cerebrovascular disease, dialysis vintage, dialysis adequacy, interdialytic weight gain, ultrafiltration volume, serum albumin, and medication use (including antiplatelet agents, vitamin K antagonists, lipid-lowering agents, β-blockers, renin–angiotensin system inhibitors, and calcium channel blockers). Model 2 included Model 1 plus mean SBP; Model 3 included Model 1 plus SBP standard deviation; Model 4 included Model 1 plus both mean SBP and SBP standard deviation. CI, confidence interval; CV, cardiovascular; HR, hazard ratio; MACE, major adverse cardiovascular events; MI, myocardial infarction; SBP-TTR, systolic blood pressure time in target range.

**Table 3.** Associations Between SBP-TTR and Cardiovascular Outcomes in Fine–Gray Competing-Risk Models.

|  | SHR (95% CI) | P value |
|---|---|---|
| MACE |  |  |
| Crude | 0.94 (0.90 - 0.99) | 0.015 |
| Model 1 | 0.92 (0.87 - 0.96) | 0.001 |
| Model 2 | 0.92 (0.88 - 0.96) | 0.001 |
| Model 3 | 0.93 (0.88 - 0.98) | 0.005 |
| Model 4 | 0.93 (0.88 - 0.98) | 0.005 |
| Nonfatal stroke |  |  |
| Crude | 0.89 (0.83 - 0.95) | <0.001 |
| Model 1 | 0.86 (0.81 - 0.92) | <0.001 |
| Model 2 | 0.87 (0.82 - 0.93) | <0.001 |
| Model 3 | 0.89 (0.83 - 0.96) | 0.002 |
| Model 4 | 0.89 (0.83 - 0.96) | 0.003 |
| CV death |  |  |
| Crude | 0.96 (0.90 - 1.02) | 0.170 |
| Model 1 | 0.94 (0.88 - 1.00) | 0.044 |
| Model 2 | 0.94 (0.88 - 1.00) | 0.041 |
| Model 3 | 0.94 (0.88 - 1.00) | 0.069 |
| Model 4 | 0.94 (0.88 - 1.00) | 0.057 |
| Nonfatal MI |  |  |
| Crude | 0.96 (0.87 - 1.06) | 0.370 |
| Model 1 | 0.93 (0.84 - 1.04) | 0.220 |
| Model 2 | 0.93 (0.83 - 1.04) | 0.200 |
| Model 3 | 0.95 (0.85 - 1.07) | 0.420 |
| Model 4 | 0.95 (0.84 - 1.07) | 0.400 |

SHRs are per 10% increment in interpolation-based SBP-TTR. Fine–Gray models accounted for competing risks, with all-cause death treated as the competing event for nonfatal outcomes and non-cardiovascular death for cardiovascular death. Model 1 included sex, age, smoking status, diabetes mellitus, cardiovascular disease, cerebrovascular disease, dialysis vintage, dialysis adequacy, interdialytic weight gain, ultrafiltration volume, serum albumin, and medication use (including antiplatelet agents, vitamin K antagonists, lipid-lowering agents, β-blockers, renin–angiotensin system inhibitors, and calcium channel blockers). Model 2 included Model 1 plus mean SBP; Model 3 included Model 1 plus SBP standard deviation; Model 4 included Model 1 plus both mean SBP and SBP standard deviation. CI, confidence interval; CV, cardiovascular; MACE, major adverse cardiovascular events; MI, myocardial infarction; SBP, systolic blood pressure; SHR, subdistribution hazard ratio; TTR, time in target range.

### Dose–response relationship and subgroup analysis

Restricted cubic spline analysis showed inverse associations of SBP-TTR with MACE (*P* for overall association = 0.019) and nonfatal stroke (*P* for overall association = 0.016), without evidence of nonlinearity (*P* for nonlinearity = 0.754 and 0.986, respectively) (**Figure 5**). Associations with CV death (*P* for overall association = 0.144) and nonfatal MI (*P* for overall association = 0.221) were not significant. Effect estimates for all four CV outcomes were directionally consistent across prespecified subgroups, with no evidence of effect modification (all *P* for interaction >0.05; **Supplementary Figure S4**).

**Figure 5.**
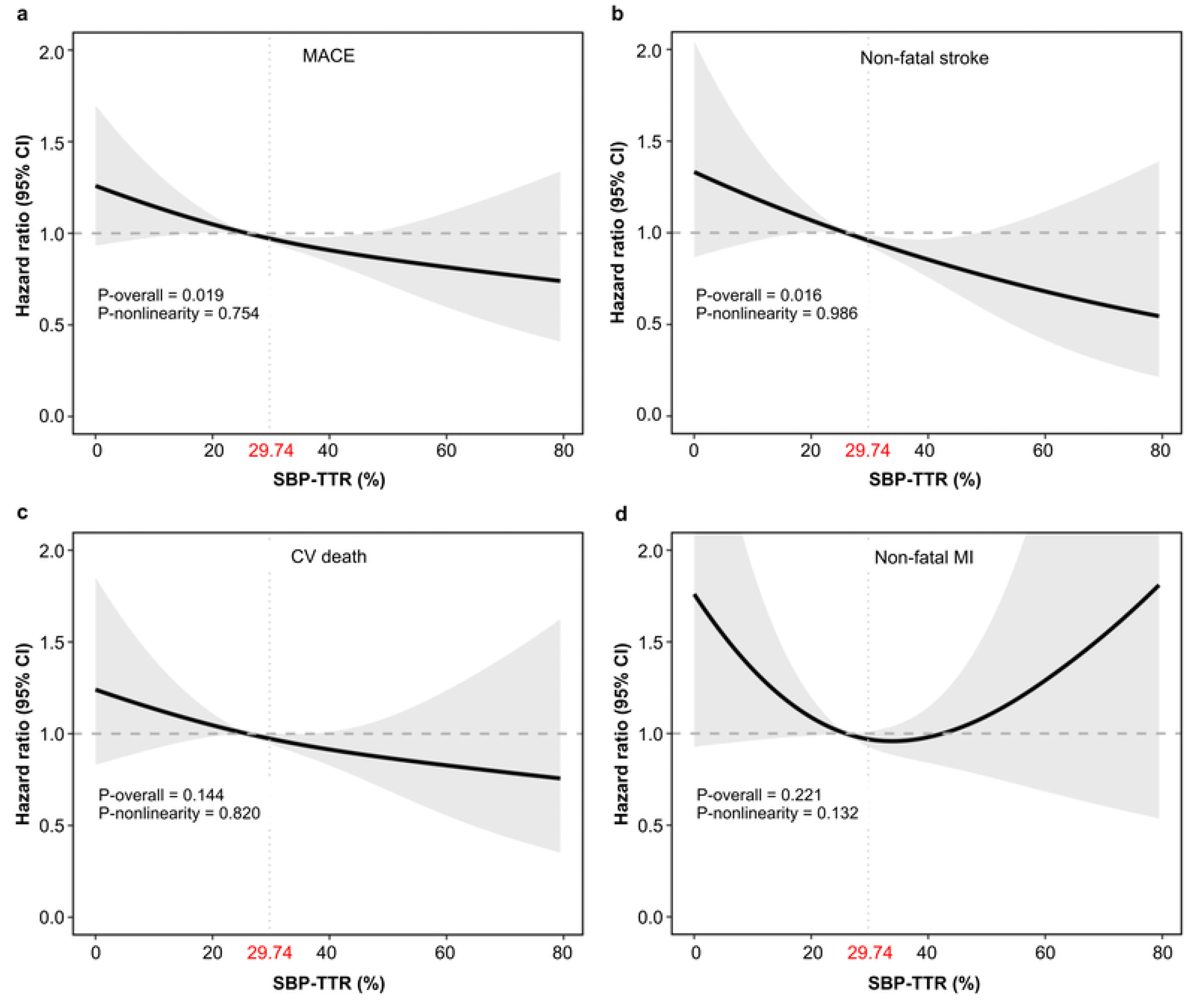
Associations Between SBP-TTR and Cardiovascular Outcomes. Restricted cubic spline models evaluated the associations of SBP-TTR with MACE (A), nonfatal stroke (B), CV death (C), and nonfatal MI (D). SBP-TTR was defined as the percentage of interpolated time during the 12-week baseline period in which pre-dialysis SBP was within 130–150 mmHg. Solid lines represent adjusted HRs, shaded areas represent 95% CIs, and horizontal dashed lines indicate an HR of 1.0. Models were adjusted for sex, age, smoking status, diabetes mellitus, CVD, cerebrovascular disease, dialysis vintage, dialysis adequacy, IDWG, UF volume, serum albumin, medication use, mean SBP, and SBP SD. P-overall indicates the overall association, and P-nonlinearity indicates departure from linearity. CI, confidence interval; CV, cardiovascular; CVD, cardiovascular disease; HR, hazard ratio; IDWG, interdialytic weight gain; MACE, major adverse cardiovascular events; MI, myocardial infarction; SBP-TTR, systolic blood pressure time in target range; SD, standard deviation; UF, ultrafiltration. Alt text: Four restricted cubic spline plots showing approximately linear inverse associations of SBP-TTR with MACE and nonfatal stroke. Associations with CV death and nonfatal MI were not statistically significant.

### Robustness across alternative definitions and analytical approaches

Results were consistent using proportion-based and dual-target SBP × DBP-TTR definitions. With the proportion-based definition, each 10-percentage-point increase was associated with lower risks of MACE (HR, 0.93; 95% CI, 0.88–0.98; *P* = 0.011) and nonfatal stroke (HR, 0.89; 95% CI, 0.82–0.97; *P* = 0.005), but not CV death or nonfatal MI **(Supplementary Table S3)**. The dual-target definition similarly identified lower risks of MACE (HR, 0.91; 95% CI, 0.84–0.99; *P* = 0.026) and nonfatal stroke (HR, 0.87; 95% CI, 0.76–0.99; *P* = 0.031) **(Supplementary Table S5)**. Exploratory outcome patterns were directionally concordant across both alternative definitions **(Supplementary Tables S4 and S6)**. Categorical analysis showed lower risks of MACE and nonfatal stroke in the highest versus lowest quartile across interpolation-based, proportion-based, and dual-target definitions **(Supplementary Tables S7, S9, and S11)**, with corresponding exploratory outcomes in **Supplementary Tables S8, S10, and S12**. Continuous Fine–Gray analysis were consistent for the proportion-based and dual-target definitions **(Supplementary Tables S13 and S14)**, and **Supplementary Table S15** summarizes all CV analysis.

### Incremental prognostic value and model refinement

Adding interpolation-based SBP-TTR to the clinical reference model improved model fit and modestly increased discrimination for MACE (C-index, 0.66 to 0.67). It also improved reclassification (NRI, 12.0%; 95% CI, 2.9–17.7) and discrimination (IDI, 1.2%; 95% CI, 0.2– 2.8), whereas adding mean SBP alone provided minimal incremental value (NRI, 0.3%; IDI, 0.1%) **(Supplementary Table S19)**. For nonfatal stroke, SBP-TTR increased the C-index from 0.68 to 0.71 and produced larger improvements in NRI and IDI. Combined models incorporating SBP-TTR and residualized BP variability yielded further gains for MACE and nonfatal stroke. In contrast, SBP-TTR alone did not improve discrimination for CV death and provided minimal incremental value for nonfatal MI; combined models produced only modest improvements. Similar qualitative patterns were observed with proportion-based and dual-target TTR definitions **(Supplementary Tables S20 and S21)**. In the primary interpolation-based analysis, the fully combined model incorporating SBP-TTR, residualized mean SBP, and residualized SBP variability yielded an NRI of 17.5% and an IDI of 1.9% for MACE. For nonfatal stroke, the C-index increased from 0.68 to 0.75, with an NRI of 30.0% and an IDI of 7.2%.

## Discussion

### Principal findings

To our knowledge, this is the first large prospective study to evaluate Rosendaal-derived SBP-TTR in patients receiving MHD. Three findings are noteworthy. First, mean pre-dialysis SBP showed U-shaped associations with MACE and nonfatal stroke, supporting 130–150 mmHg as a clinically interpretable range for longitudinal assessment. Second, higher SBP-TTR within this range was independently associated with lower risks of MACE and nonfatal stroke after adjustment for mean SBP and visit-to-visit variability; each 10-percentage-point increase corresponded to 7% and 11% lower risks, respectively. Associations were consistent in Fine–Gray models, categorical analysis, and alternative TTR definitions. Third, SBP-TTR improved model fit and risk reclassification beyond conventional BP metrics, whereas mean SBP alone provided little incremental value. For MACE, adding interpolation-based SBP-TTR increased the C-index from 0.66 to 0.67 and yielded an NRI of 12.0% and IDI of 1.2%; gains were larger for nonfatal stroke. These findings suggest that CV risk in HD reflects not only average BP but also the consistency with which BP remains within a clinically relevant range.

### Identification of a clinically relevant pre-dialysis SBP range

The 130–150 mmHg range accords with prior reports of U-shaped associations between dialysis-unit BP and adverse outcomes.^15^ Intensive single-value targets derived from non-dialysis populations may not apply directly to HD.^16^ In the HEMO study, pre-dialysis SBP <120 mmHg was associated with higher mortality than 140–159 mmHg,^17^ while CRIC identified the lowest CV risk between 140 and 170 mmHg.^18^ Jhee et al. similarly observed increased mortality at both <110 and ≥170 mmHg.^19^ The BID Pilot Study found no CV benefit from targeting 110–140 mmHg and reported more intradialytic hypotension and vascular-access thrombosis,^20,21^ and systematic reviews have not established a net benefit of lower targets.^22,23^ In HD, low pre-dialysis SBP may reflect impaired perfusion, excessive UF, or limited cardiac reserve, whereas high SBP may indicate volume overload and arterial stiffness. Thus, 130–150 mmHg should be viewed as a plausible analytical window for assessing longitudinal stability rather than as a treatment target. Prospective validation is required before clinical implementation.

### Interpretation of SBP-TTR in HD

SBP-TTR integrates the magnitude and persistence of BP control and therefore complements mean SBP and variability measures.^9,11,24^ Mean SBP summarizes overall exposure but obscures excursions outside a clinically relevant range; variability measures quantify dispersion without indicating whether readings are favorable. TTR captures both dimensions in a time-weighted metric. This distinction is particularly relevant in HD, where pre-dialysis BP is repeatedly altered by interdialytic fluid accumulation, UF volume, arterial stiffness, autonomic dysfunction, and limited cardiac reserve. In this cohort, mean SBP displayed a nonlinear risk pattern, whereas higher SBP-TTR was associated with progressively lower risk without evidence of nonlinearity, translating a complex BP–risk relationship into a clinically interpretable longitudinal measure.

SBP-TTR probably reflects more than pharmacological control. Low values may indicate unstable volume management, UF intolerance, treatment patterns, arterial stiffness, frailty, poor intake, or limited CV reserve. Interdialytic SBP rises with weight gain, and the BP response to volume accumulation is modified by ventricular and arterial stiffness.^25,26^ SBP-TTR may therefore represent an integrated hemodynamic phenotype encompassing volume status, ventricular–arterial coupling, UF tolerance, myocardial reserve, and treatment practices. The observation that mean SBP was similar across TTR quartiles, while variability and outcomes differed, further suggests that averages alone do not fully characterize CV burden. This is consistent with ND studies showing prognostic value of TTR beyond mean BP^11,27^ and with dialysis studies linking BP variability to adverse outcomes.^28,29^ Residualization analysis likewise indicated that SBP-TTR contained information not explained by mean SBP or variability, supporting its potential value as a complementary rather than substitutive BP metric.

### Outcome-specific associations

Associations were strongest for nonfatal stroke, particularly ischemic stroke and large-artery atherosclerotic disease, but were not significant for nonfatal MI or CV death. Cerebral circulation may be especially vulnerable to recurrent perfusion-pressure fluctuations during HD because of intradialytic hemodynamic stress and impaired autoregulation.^30,31^ Repeated instability may promote intermittent cerebral hypoperfusion, endothelial injury, and progression of large-vessel disease.^32^ By contrast, nonfatal MI and CV death reflect heterogeneous mechanisms, including arrhythmia, myocardial stunning, structural heart disease, and hemodynamic overload,^33–35^ which may not be captured by pre-dialysis SBP-TTR alone. This outcome-specific pattern suggests that SBP-TTR represents one component of the broader CV risk architecture of HD rather than a global surrogate for all CV events.

### Threshold effects and dose–response effect of SBP-TTR

Spline analysis revealed that CV risk began to decline once SBP-TTR exceeded approximately 30%, with further increases associated with progressively lower risk and no clear plateau. This pattern contrasts with the U-shaped associations typically observed for mean SBP and with higher TTR thresholds reported in general hypertensive populations, where optimal cardioprotection often requires SBP-TTR > 65%.^11,27,36^ Although exploratory, these thresholds provide a clinically intuitive framework for interpreting longitudinal BP stability in HD.

### Clinical implications

SBP-TTR can nevertheless be calculated from routinely collected pre-dialysis measurements without additional devices and may support periodic hemodynamic assessment. A low or declining SBP-TTR could prompt review of dry weight, interdialytic sodium and fluid intake, UF tolerance, antihypertensive treatment, intradialytic symptoms, and dialysis-day medication practices. Serial calculation across successive 12-week periods may help identify changes in hemodynamic stability even when mean SBP remains unchanged. However, SBP-TTR should be interpreted alongside volume status, IDWG, cardiac function, and susceptibility to intradialytic hypotension rather than used as a standalone treatment target.

### Limitations

Several limitations merit consideration. First, the observational design precludes causal inference, although the prespecified 12-week exposure window preceded outcome follow-up and competing-risk models addressed the high mortality rate. Second, residual confounding remains possible, particularly from medication adherence, dietary sodium intake, frailty, nutritional status, and healthcare engagement. The gradient in lipid-lowering therapy across SBP-TTR quartiles may indicate differences in care or adherence despite extensive adjustment. Third, exposure was based on dialysis-unit pre-dialysis BP and did not include home or ambulatory measurements, which may better represent interdialytic BP burden. The time-weighted approach and consistency across alternative TTR definitions improve robustness but do not establish equivalence with out-of-unit BP. Fourth, both the 130–150 mmHg range and quartile boundaries were data-informed, introducing potential overfitting and circular inference because outcome patterns contributed to exposure definition. Although multiple candidate ranges and sensitivity analysis were examined, independent validation is essential. Fifth, residualization assumed linear relationships among BP metrics, and the NRI, IDI, and discrimination estimates may be optimistic when derived and evaluated in the same cohort. Finally, the cohort was drawn from dialysis units in one region of Taiwan, potentially limiting generalizability. Independent multinational validation and prospective studies are needed before SBP-TTR or the proposed range informs clinical targets.

### Conclusions

Among patients undergoing maintenance hemodialysis, higher SBP-TTR within 130– 150 mmHg was associated with lower risks of nonfatal stroke and major adverse cardiovascular events, independent of mean SBP and SBP variability. SBP-TTR may complement conventional blood pressure measures for cerebrovascular and cardiovascular risk assessment, but external validation is required before clinical implementation.

## Data Availability

Data can be obtained upon a reasonable request.

## Acknowledgments

The authors thank Enago for English-language editing assistance.

## Author Contributions

Fan-Yu Chen: Conceptualization, Data curation, Formal analysis, Investigation, Methodology, Project administration, Software, Validation, Visualization, Writing – original draft, Writing – review & editing.

Mu-Yang Hsieh: Conceptualization, Investigation, Methodology, Resources, Validation, Writing – review & editing.

Zih-Kai Kao: Data curation, Software, Validation, Visualization, Writing – review & editing. Shao-Yuan Chuang: Formal analysis, Methodology, Software, Validation, Writing – review & editing.

Ru-Yin Shu: Data curation, Investigation, Resources, Validation, Writing – review & editing.

Chen-Huan Chen: Conceptualization, Methodology, Supervision, Validation, Writing – review & editing.

Mei-Jy Jeng: Conceptualization, Funding acquisition, Methodology, Project administration, Resources, Supervision, Validation, Writing – review & editing.

Chih-Cheng Wu: Conceptualization, Funding acquisition, Methodology, Project administration, Resources, Supervision, Validation, Writing – review & editing.

Hao-Min Cheng: Conceptualization, Funding acquisition, Methodology, Project administration, Resources, Supervision, Validation, Writing – review & editing.

All authors reviewed and approved the final version of the manuscript and agreed to be accountable for all aspects of the work.

## Generative Artificial Intelligence Disclosure

During the preparation of this work, the authors used ChatGPT (OpenAI) for language editing, manuscript organization, and formatting. After using this tool, the authors reviewed and edited the content as needed and take full responsibility for the content of the manuscript.

## Abbreviations and Acronyms

AF: atrial fibrillation
ARV: average real variability
BP: blood pressure
CI: confidence interval
CV: cardiovascular
CVD: cardiovascular disease
DBP: diastolic blood pressure
ESRD: end-stage renal disease
FDR: false discovery rate
HD: hemodialysis
HF: heart failure
HR: hazard ratio
IDI: integrated discrimination improvement
IDWG: interdialytic weight gain
MACE: major adverse cardiovascular events
MHD: maintenance hemodialysis
MI: myocardial infarction
ND: nondialysis
NRI: net reclassification improvement
SBP: systolic blood pressure
SBP-TTR: systolic blood pressure time in target range
SD: standard deviation
SHR: subdistribution hazard ratio
TTR: time in target range
UF: ultrafiltration
VA: vascular access.

